# Clinically meaningful metrics of speech in neurodegenerative disease: Quantification of speech intelligibility and naturalness in ataxia

**DOI:** 10.1101/2023.03.28.23287878

**Authors:** Adam P. Vogel, Paul Maruff, Hannah Reece, Hannah Carter, Geneieve Tai, Benjamin G. Schultz, Louise Corben, Martin B Delatycki, Athanasios Tsanas

## Abstract

**Background:** Digital biomarkers continue to make headway in the clinic and clinical trials for neurological conditions. Speech is a domain with great promise.

**Objectives:** Using Friedreich ataxia (FRDA) as an exemplar population, we aimed to align objective measures of speech with markers of disease severity, speech related quality of life and subjective judgements of speech using supervised machine learning techniques.

**Methods:** 132 participants with genetically confirmed diagnosis of FRDA were assessed using digital speech tests, disease severity scores (Friedreich Ataxia Rating Scale, FARS) and speech related quality of life ratings over a 10-year period. Speech was analyzed perceptually by expert listeners for intelligibility (ability to be understood) and naturalness (deviance from healthy norm) and acoustically across 344 features. Features were selected and presented into a random forest and a support vector machine classifier in a standard supervised learning setup designed to replicate expert-produced scores.

**Results:** We demonstrate a subset of measures are strongly associated with all four clinical scales. Objective speech data replicated experts’ assessments of naturalness and intelligibility. These scores represent a lower level of variability than observed between subjective listener ratings. Findings provide evidence there are specific objective markers of speech that change over time and reflect clinical aspects of the disease.

**Discussion:** The use of a large dataset yielded a speech assay capable of accurately approximating expert listener ratings of key clinical aspects of dysarthria severity. Distinct but complementary subsets align with disease severity and speech related quality of life.

## Introduction

Friedreich ataxia (FRDA) is an autosomal recessive multisystem neurodegenerative disorder caused most commonly by homozygosity for a GAA triplet repeat expansion in intron 1 of the *FXN* gene leading to reduced frataxin, a protein important for mitochondrial iron metabolism ^1^. FRDA presents with clear and progressive dysarthria, even in early disease stages, making speech assessment central to diagnosis and management. The ability to clearly communicate ideas and feelings is central to social acts in healthy adults and has great importance in individuals whose neurologic disease progressively increases limitation for daily activities. Consequently, the dysarthria that characterizes FRDA is associated with a substantial reduction in quality of life (QoL) and remains an important target for speech behavioral therapies and for curative pharmacological therapies. Clear models of dysarthria are therefore important for both diagnoses and management of FRDA.

Expert assessment of clinical speech in FRDA shows the specific dimensions of dysarthria manifest as two overarching dimensions of communication difficulty that act to reduce speech-related QOL. First, there is a reduction in the listener’ s capacity to understand what is being said, a dimension termed intelligibility. In FRDA reduced intelligibility leads to feelings of isolation, reduced self-esteem, frustration, difficulty talking to people unfamiliar with the speaker. Second, in adults with FRDA speech differs from that expected from healthy individuals, a dimension termed naturalness. In FRDA, a reduced naturalness in speech often leads listeners to assume the presence of alcohol intoxication or cognitive impairment^2^. Currently assessment of these dimensions of speech-related QOL requires expert listeners make judgements about acoustic characteristics of spontaneous and cued expression in response to standardized questions and commands. While providing a strong basis for diagnoses, the use of subjective scoring on perceptual scales reduces their consistency, within- and between-clinics, and even within-assessors ^3^ thereby limiting the potential for objective assessment of speech to be applied widely in decisions about the identification, treatment and management of FRDA.

Recent improvements in digital technology, including microphone design, computational power and signal processing now provide effective and usable tools for both collection and analyses of speech samples with a fidelity sufficient for objective analyses. Information arising from use of digital assessment of expressive speech have guided decisions about the presence ^4-6^ and progression ^7^ of brain diseases, as well as about their response to behavioral and pharmacotherapies ^8, 9^. However, as for all aspects of complex clinical information shifts from expert human assessment to digital measurement can result in loss of clinical relevance ^10^. For feature rich data such as expressive speech, such loss can occur because information is reduced to highly specific but theoretically constrained characteristics (e.g., periodicity of vocal fold closure) that while sensitive to impairment have low clinical relevance. Alternatively data science approaches taken to optimize expressive speed data sets provide aggregate outcomes that optimize variance but have no clinical meaning ^16,17^. One way to overcome these limitations in FRDA would be to constrain data science analyses of digitally collected speech samples by characteristics, such as intelligibility and naturalness, that have established clinical relevance to the disease. The aim of this study was to explore the potential for quantification of speech intelligibility and naturalness, data was collated across a 10-year period for a large cohort of individuals with dysarthria associated with FRDA. Digital speech data were compared to corresponding clinical speech assessments as well as clinician rated measures of intelligibility and naturalness and standardized patient reported speech quality of life scores.

## Methods

### Data

Data were acquired from 132 patients with FRDA (62 males), with age at first visit (median±iqr) 31±18.5 years. Of those 132 participants, 36 had a single visit, 22 two visits, 16 had three visits, and the remaining 58 participants had four or more visits over ten years. Data from 36 of these participants was published previously ^11^. Participants completed the following speech tasks: (a) sustained vowel /a/ phonation, (b) monologue, (c) reading a set paragraph, (d) days of the week, and (e) counting numbers 1 to 20. For each visit we had 1-6 sustained vowel /a/ phonations (for most participants 2 or 3 sustained vowels per visit). In addition to those tasks, participants were assessed using standardized clinical scales assessing FRDA and speech, the Friedreich ataxia rating scale (FARS) ^12^, and the speech subscale of the Friedreich ataxia impact scale (FAIS) ^13^. The study was approved by The University of Melbourne Human Research Ethics Committee (#1750022) and all participants provided written informed consent.

Two expert listeners (5+ years of experience) blinded to disease features rated all monologue and vowel samples for *intelligibility* and *naturalness*. Both measures were rated on a five-point scale where 0= unremarkable, 1=sub-clinical, 2= mild, 3= moderate and 4= severe, according to methods published previously ^7, 14, 15^. Raters provided an individual score. Where there was disagreement, raters produced an agreed consensus score. *Inter-rater variability* was assessed of the two experts. Findings were summarized in the form of agreement matrices (similarly to confusion matrices). Entries in the long diagonal of Tables S1 and S2 denote agreement between experts, entries off the long diagonal indicate the differences observed between the two experts. For speech related quality of life measures, scores from the speech subscale of the FAIS (1 of 8 scales) were used. FAIS subscale scores were transformed to have a range of 0 to 100, with a higher score indicating greater impact of FRDA on speech. FARS is a clinical rating scale administered by an experienced physician. The total FARS score was used which has range of 0 to 159 with higher scores denoting increased disease severity.

Overall, in terms of unique patient visits, 468 sustained vowel /a/ phonations, 253 recordings with the reading passage, 302 monologue recordings, 197 recordings with days of the week, and 133 recordings with counting were processed. Timing and relative number of assessments for participants in the study is illustrated in Figure 1. Corresponding values for 419 of those samples for naturalness and intelligibility, 336 samples matched with FARS, and 246 samples matched with FAIS were obtained. We used the k-Nearest Neighbor (kNN) approach ^16^ to account for missing data (see Supplementary materials for detail on data imputation)

**Fig. 1:**
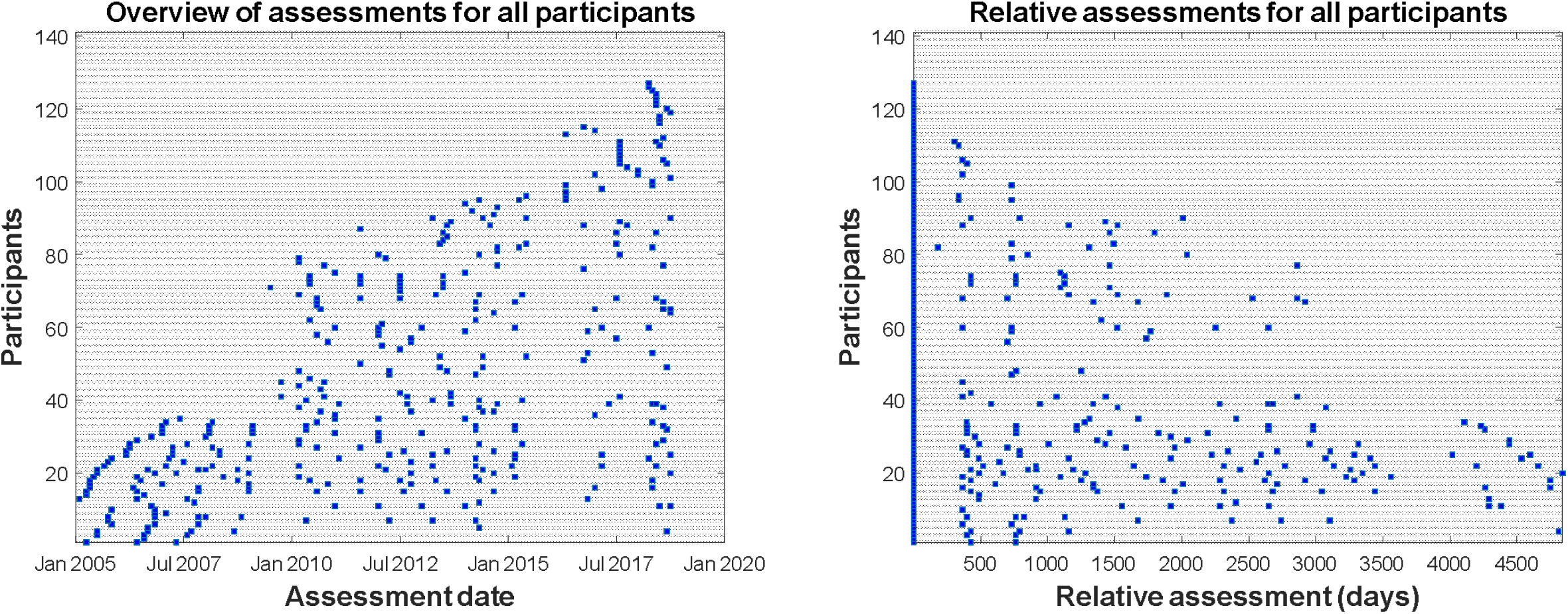
Timing and relative number of assessments for participants in the study.

### Acoustic analysis

Across the four connected speech samples (i.e., days of the week, counting, reading and monologue), we applied a range of 9 broad measures of *timing* described elsewhere ^14, 17, 18^, which we refer to as *timing measures* (4×9=36 used here). These include measures of mean pause length, percentage of silence in a sample, speech rate and variability of pause length. For the sustained vowel /a/ phonations, we applied a range of 308 *dysphonia measures* previously described in detail ^19, 20^ and in Supplementary materials. These dysphonia measures capture a range of vocal problems from traditional *jitter measures* (variability in fundamental frequency), *shimmer measures* (variability in signal amplitude), Mel-Frequency Cepstral Coefficients (MFCCs), to more advanced nonlinear measures such as Vocal Fold Excitation Ratio (VFER) and other energy- and entropy-based algorithmic approaches.

In total, we summarized all the extracted measures to form a design matrix 434 × 344 (308 dysphonia measures + 36 timing measures). For simplicity, we will refer collectively to all 344 extracted speech signal characteristics as *acoustic measures*. To adhere to the machine learning terminology, when these (or a subset of those) acoustic measures are presented in the subsequent statistical mapping phase, they are referred to as *features*. In all cases, the aim was to associate and develop a functional mapping of the acoustic measures to the two clinical scales (i) FARS, (ii) FAIS, and two expert rater outcomes (iii) naturalness and (iv) intelligibility. Finally, we aimed to assess whether the features used were sensitive to longitudinal clinical outcome changes.

### Quantizing FARS and FAIS

In addition to working with the provided FARS and FAIS scores, we used quantized versions (constrained set of integers rather than large multi-feature dataset). This is because the two clinical scales span a large range, and we have a relatively small number of samples. We also wanted to investigate if speech measures could predict ranges rather than the original FARS and FAIS score itself. We used a simple uniform quantization with 10 levels, i.e., FARS and FAIS were mapped separately onto 10 levels with the aim to replicate those levels using the acoustic features.

### Statistical analysis

Spearman correlation coefficient was used to quantify the statistical strength of the associations, using the empirical rule of thumb that these associations are deemed *statistically strong* when the magnitude of the correlation was above 0.3, i.e. |R| > 0.3 ^21^. We assessed a statistical relationship as *statistically significant* at the p=0.01 level.

### Statistical mapping

Random Forests (RF) were used for statistical mapping of the acoustic measures to estimate each of the four outcomes (two clinical scales, two listener ratings). The inherent Feature Selection (FS) was determined as part of the Cross Validation (CV) process following the methodology to select the final feature set using the voting approach previously described ^20^.

### Model validation and performance metrics

Model performance was assessed using 10-fold CV with 100 iterations for statistical confidence ^16^, similar to our previous work ^5, 19, 20^. All four outcomes were treated as standard classification settings, and for convenience the Mean Absolute Error (MAE) was used to assess performance summarized on a single scalar number (rather than e.g., confusion matrices). In all cases we reported results using mean and standard deviation to present the uncertainty around the quoted mean error in the out-of-sample computations. For the quantized FARS and FAIS, confusion matrices are provided to illustrate the performance of the algorithms and determine which classes (quantized levels) might be confusing for the classifier (this makes it easier to identify problem with model estimates for the 10 quantized levels).

### Feature transformation using Principal Component Analysis (PCA)

Principal Component Analysis (PCA) ^22^ was used to obtain an aggregate (collective) approach that combined the individual features. PCA linearly projects the original features onto principal components, which aim to minimize the explained variance in the dataset. We computed the first three principal components, aiming to investigate how the variability of those three principal components change as a function of time, to directly compare these aggregate scores against the longitudinal changes of the clinical scales.

## Results

Table 1 summarizes the 10 most strongly correlated features with each of the four clinical outcomes. The speech phenotype of FRDA is dominated by timing deficits rather than dysphonia, as evidenced in Table 1. How the timing measures were statistically associated with each other was explored using a colored correlation matrix (correlogram, see Supplementary Materials in Fig. S1).

**Table 1:** Statistical associations using the Spearman correlation coefficient between the features and each of the clinical outcomes.

| FARS |  | FAIS |  | Naturalness |  | Intelligibility |  |
| --- | --- | --- | --- | --- | --- | --- | --- |
| Feature | R | Feature | R | Feature | R | Feature | R |
| 'Days dur' | 0.572 | 'Days ssum' | 0.355 | 'READ pmean' | 0.668 | 'READ pmean' | 0.657 |
| 'Days srate' | -0.572 | 'READ_Flux_stdPauseDur' | 0.343 | 'READ_Energy_meanSylPeriod' | 0.652 | 'READ_Energy_meanSylPeriod' | 0.606 |
| 'READ spratio' | -0.509 | 'det_TKEO_mean_5_coef' | 0.327 | 'READ_Energy_SylsPerSec' | -0.652 | 'READ_Energy_SylsPerSec' | -0.606 |
| 'READ pperc' | 0.480 | 'READ_Flux_stdSylPeriod' | 0.324 | 'READ_Energy_meanPauseDur' | 0.603 | 'READpstdev' | 0.560 |
| 'READ_Energy_meanSylPeriod' | 0.467 | 'det_entropy_shannon_5_coef' | -0.317 | 'READpstdev' | 0.599 | 'READ_Energy_meanPauseDur' | 0.556 |
| 'READ_Energy_SylsPerSec' | -0.467 | 'det_TKEO_std_5_coef' | 0.316 | 'READ_Energy_stdSylPeriod' | 0.589 | 'READ_Energy_stdSylPeriod' | 0.550 |
| 'READ_Energy_SpeakPercent' | -0.463 | 'Jitter->F0_TKEO_prc5' | -0.315 | 'READ spratio' | -0.559 | 'READ spratio' | -0.549 |
| 'READ_Energy_PausePercent' | 0.463 | 'READ_Energy_SpeakPercent' | -0.313 | 'READ_Energy_stdPauseDur' | 0.551 | 'READ_Energy_stdPauseDur' | 0.504 |
| 'READ_Energy_Speech2PauseRatio' | -0.463 | 'READ_Energy_PausePercent' | 0.313 | 'READ pperc' | 0.521 | 'READ pperc' | 0.484 |
| 'Jitter->F0_abs0th_perturb' | 0.454 | 'READ_Energy_Speech2PauseRatio' | -0.313 | 'READ_Energy_SpeakPercent' | -0.481 | 'READ dur' | 0.438 |

For simplicity we only present the top 10 most strongly correlated features with each of the four outcomes. We used all 344 features to determine the strongest statistical associations. All relationships above were statistically significant ( $p < 0.0001$ ). READ = reading passage; DAYS = days of the week.

Given the measures extracted from the reading passage dominate the naturalness and intelligibility outcomes, we explored internal pairwise relationships (see Fig. S2). This was a crucial step to understand how well internally the features extracted from the reading passage were correlated with one another. Fig. S2 focuses on the features for the reading passage that were presented in Fig. S1, also presenting scatter plots to better observe outcomes. Fig. S3 is a histogram with the distributions for the four clinical scales used in the study. Table 2 presents the features selected using RF for each of the four problems investigated here.

**Table 2:** Selected feature subset using Random Forests (RF) for each of the four problems.

| **FARS** | **FAIS** | **Naturalness** | **Intelligibility** |
| --- | --- | --- | --- |
| MFCC_9th coef | MFCC_9th coef | READ_Energy_SylsPerSec | READ_pmean |
| IMF->NSR_TKEO | IMF->NSR_TKEO | IMF->NSR_TKEO | READ_Energy_meanPauseDur |
| MFCC_2nd coef | 2nd delta-delta | READ_pmean | READ_Energy_stdSylPeriod |
| MFCC_7th coef | MFCC_2nd coef | Days srate | Shimmer->Ampl_PQ3_classical_Baken |
| Jitter->pitch_PQ5_classical_Baken | det_entropy_shannon_1_coef | READ spratio | READ_Energy_meanSylPeriod |
| det_entropy_shannon_1_coef | MFCC_3rd coef | READ_Energy_meanSylPeriod | READ_Energy_SylsPerSec |
| 4th delta-delta | MFCC_7th coef | READ_Energy_meanPauseDur | READ_Flux_stdSylPeriod |
| MFCC_3rd coef | MFCC_4th coef | Days pstdev | READpstdev |
| 9th delta-delta | 4th delta-delta | READ_Flux_SylsPerSec | MONO speech pmean |
| READ_Energy_SpeakPercent | Jitter->pitch_PQ5_classical_Baken | IMF->SNR_SEO | MONO speech psddev |
| 15.45±2.88 | 15.01±3.99 | 0.30±0.08 | 0.40±0.11 |

For simplicity only the top 10 selected features for each problem are presented in descending order of RF feature importance. The last row indicates the out of sample performance when presenting the classifier with those ten features. The results are obtained using 10-fold cross validation with 100 iterations for statistical confidence. For the FARS and FAIS estimation the RF was presented only with the dysphonia measures because this led to better results than presenting the entire feature set. READ = reading passage; MONO = monologue

Fig. 2 presents the out of sample MAE for intelligibility (2.a) and naturalness (2.b). In both cases the MAE was relatively small, suggesting a robust subset with <20 features is sufficient to automatically replicate the clinical scales of naturalness and intelligibility.

**Fig. 2:**
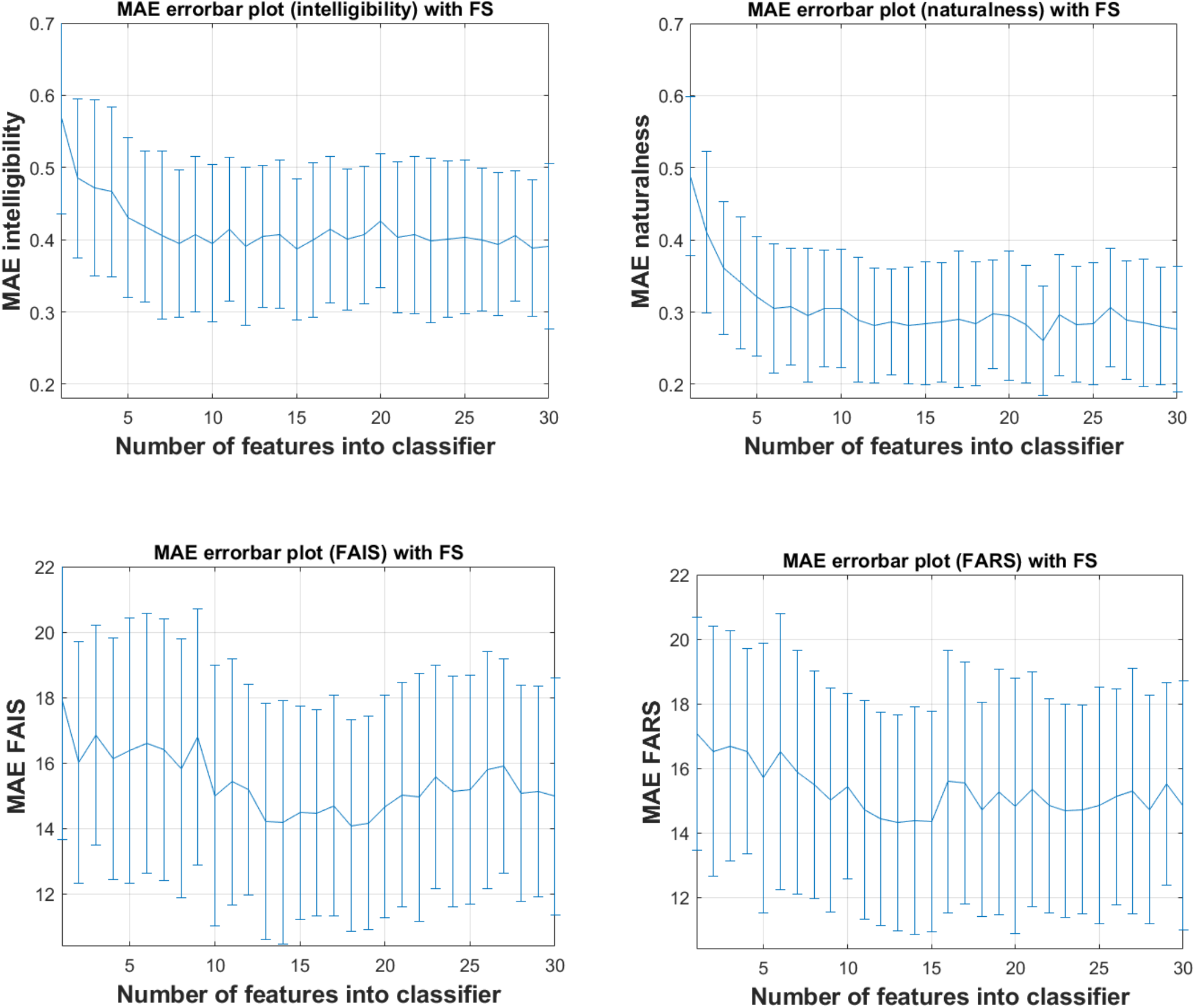
Out of sample performance for assessing a) intelligibility, b) naturalness, c) FAIS, d) FARS. Mean Absolute Error (MAE) was computed using 10-fold CV with 100 iterations for statistical confidence. Error bars represent the standard deviation around the quoted mean. FS = Feature Selection.

### Estimating FARS and FAIS using quantized versions with 10 levels

The relatively limited number of samples combined with the large deviation of the ground truth (clinicians’ estimates) and the RF estimates for FARS and FAIS motivated the need to explore quantizing the full span of the two clinical scales into a more manageable multi-class classification problem using quantization levels: both were uniformly quantized using 10 levels. Table 3 presents the out of sample results using 10-fold CV with 100 iterations when training and testing on those 10 levels for FARS and FAIS. Ideally, the long diagonal dominates and all entries off the long diagonal are zero for those two confusion matrices. Overall, the true classes are accurately determined (most entries are in the long diagonal). Using the ten levels, the MAE for FARS is (mean±std) 0.98±0.24, and for FAIS 1.34±0.37. This suggests that overall speech can accurately lead to the estimation of approximate FAIS and FARS scores, even though based on the available data the full granularity of the two metrics are not adequately replicated.

**Table 3:**
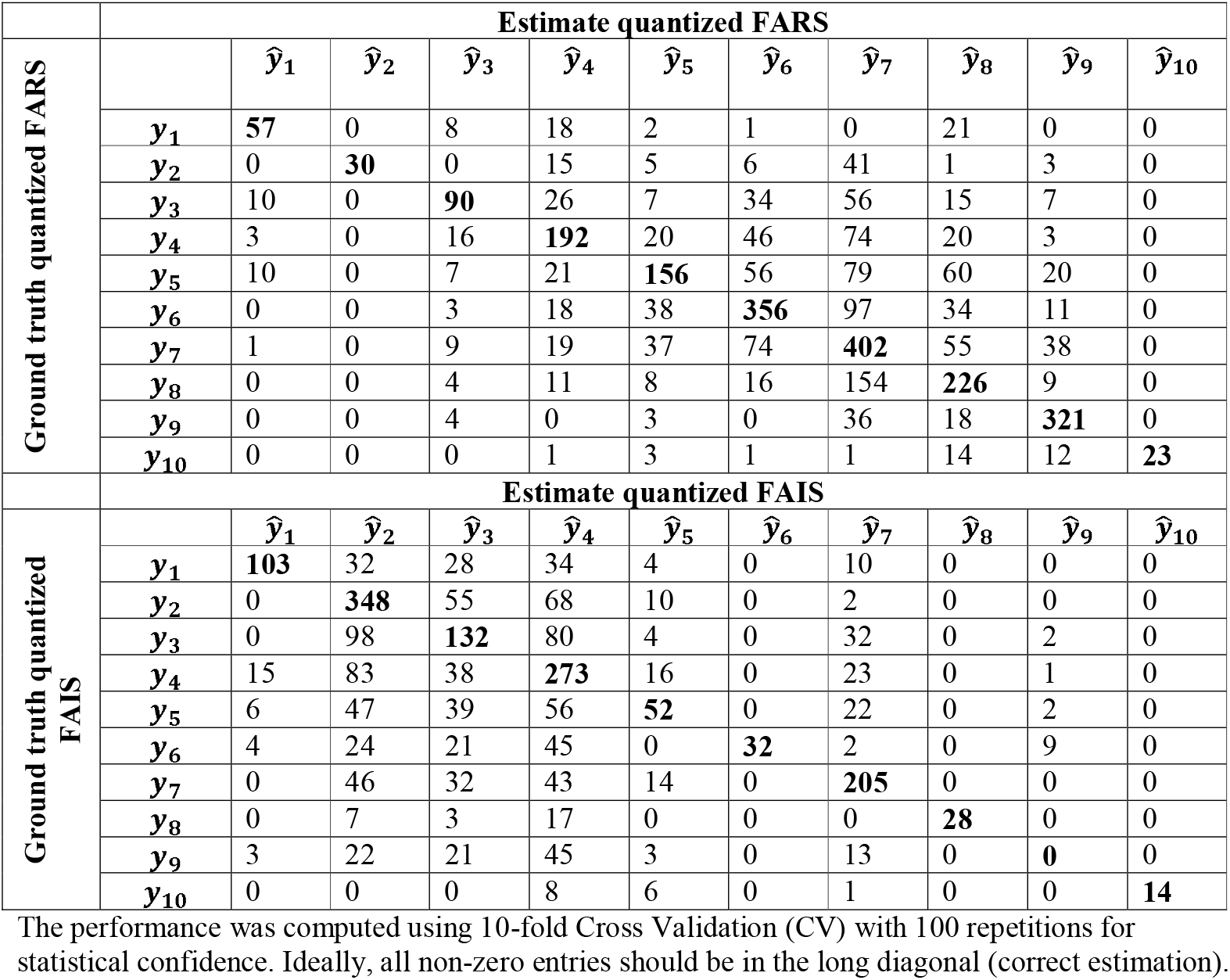
Confusion matrix presenting the out of sample performance of Random Forests (RF) to estimate the 10 quantized levels of FARS and FAIS.

### Exploring inter-rater variability for speech assessment

Table S1 summarizes the differences in clinical scoring of intelligibility between the two trained experts, and S2 summarizes the differences in clinical scoring of naturalness. For the intelligibility scores, experts exactly agreed on 266 out of the 380 samples (70% agreement), whereas for the naturalness scoring, experts agreed on 233 out of the 380 samples (61.3% agreement). This can be expressed in terms of the mean absolute difference between the two raters, which provides a guide to overall agreement between experts (which can be compared against the accuracy of the automated statistical mapping algorithms). This is effectively identical to how MAE was computed between the consensus and the automatically determined labels using statistical learning algorithms. For intelligibility, the mean absolute difference between the two raters was 0.39, and for naturalness the mean absolute difference between the two raters was 0.30. Importantly, when applying RF to estimate intelligibility and naturalness (Fig. 2), the MAE is *lower* than the inter-rater variability.

### Exploring longitudinal voice changes

Figure 3 presents longitudinal changes in intelligibility and naturalness, along with the corresponding aggregate acoustic information using the first three principal components. These principal components appear sensitive to the trajectory of the observed longitudinal changes and may be useful as markers to guide clinical decisions on longitudinal changes. This may be particularly important as in the examples above where there are marked deviations between the two expert raters.

**Fig. 3:**
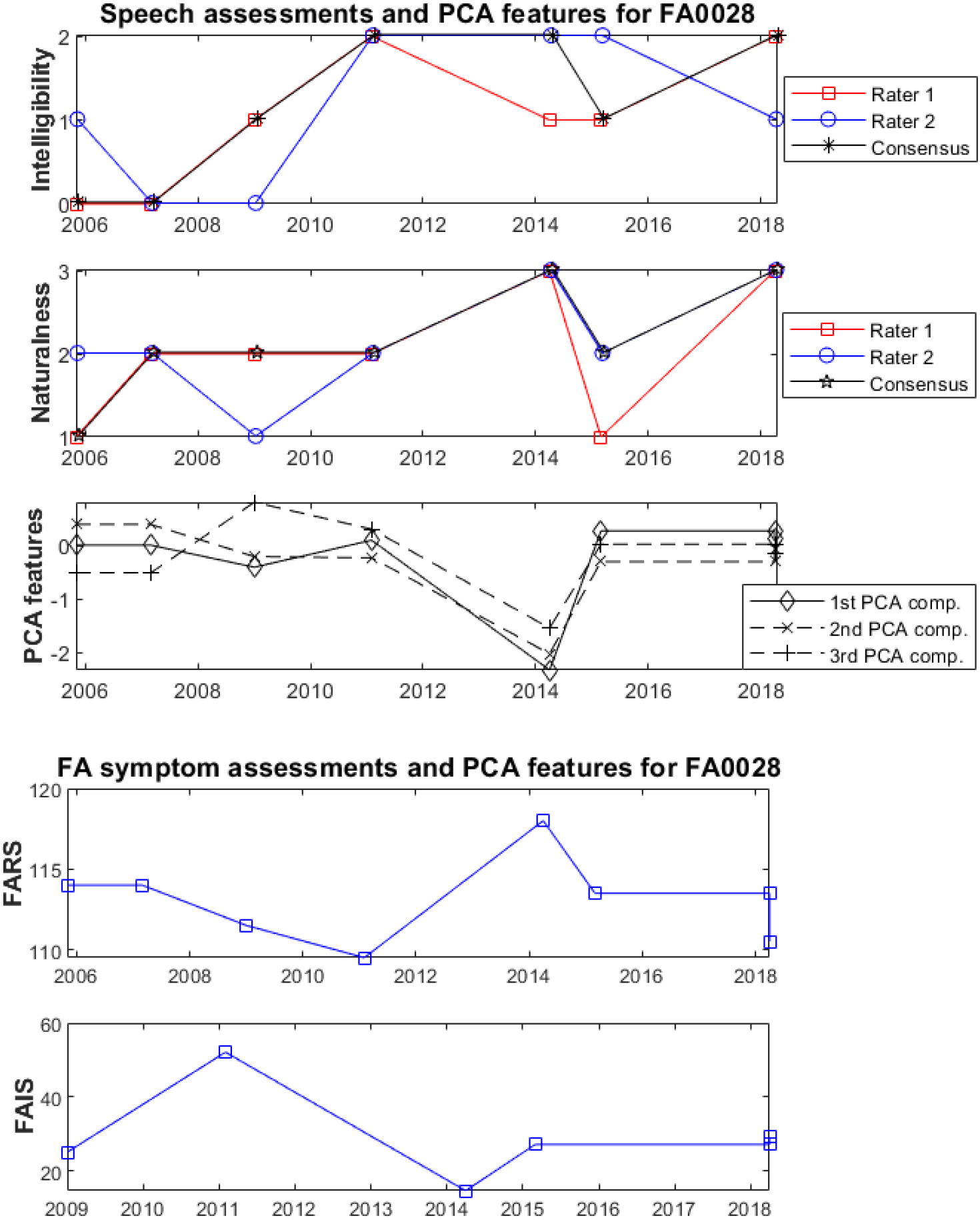
Longitudinal variability of the four scales for participants who had at least four visits (although the scales were not necessarily available for all their visits). Indicative examples of longitudinal variation of intelligibility and naturalness and the aggregate acoustic information presented using the first three principal components. a) intelligibility, b) naturalness, c) PCA features, d) FARS, e) FAIS. Note: To facilitate visualization we have introduced slight jitter on the x-axis so that the markers do not completely overlap at the time of assessment. PCA feature subset based on voice measures only and did not include measures of timing due to inadequate tokens.

## Discussion

We investigated the potential of using objective speech modalities to replicate standardized, clinician rated naturalness and intelligibility and disease specific clinical scales (FARS, FAIS) which reflect symptom severity and patient response to disease including QOL. The MAE for predicting intelligibility and naturalness in ataxia was 0.23 and 0.27 respectively for out of sample assessments. These values are lower than the variability observed between expert raters. The speech related QOL score was associated with a combination of timing and dysphonia measures whereas the indices of both intelligibility and naturalness were described by a subset of timing measures derived from automated and reading tasks. Attempts to align multi-feature speech subsets with overall disease severity scores and self-reported QOL measures yielded the largest error rates. The link between speech and overall disease is clear.

We advocate that speech is a meaningful domain itself, forming part of a battery of measures that can describe disease progression or treatment response. To mechanize this approach, consideration of how we can best describe speech is needed so that outcomes make sense to the user (either patient, clinician or trialist) and provide useful data for monitoring performance. Intelligibility and naturalness are two comprehensive features that capture important aspects of the phenotype, the speakers’ ability to be understood, and deviation from expectations about healthy expression. Changes to either of these domains can have profound impacts on the speaker, reducing independence and psycho-social well-being, producing social marginalization, triggering altered self-identity ^23, 24^ and impede or prevent both social and professional interactions leading to daily disadvantage, and underemployment2.

The raw FARS and FAIS scores were not estimated to the same level of accuracy as were outcomes based on speech alone. This could be because (a) FARS measures reflect the disease as a whole, with speech measured as a sub-component of the tool, (b) the FAIS is a subsection of a larger questionnaire that the subjectivity in FARS and FAIS assessments and their inherent variability is not captured by the speech modalities used in the study; (c) that the applied methodology with the extracted features and the machine learning methodology needs to be further refined; (d) the statistical power of samples is limited towards estimating these outcomes at the granularity they have been described. On the last point, we explored quantized levels of FARS and FAIS with the aim to replicate those quantized levels using the computed features and were able to successfully replicate the 10 quantized FARS and FAIS levels to varying degrees. It is reasonable to expect a stronger correlation between speech scores and the FAIS subscale (which is specific to speech) than the FARS in which speech is a small component. However, this difference did not manifest here, perhaps due to the inherent variability in Patient Reported Outcomes or tools like FAIS that are not designed and validated to specifically to measure speech related quality of life.

Speech is useful for mapping closely to disease severity scales, but it is better represented as its own domain warranting separate consideration. Our data suggests that speech working as a multifactorial subset of features can provide an estimate of function within a single decile, and thereby is linked but not wholly driven by disease severity.

The ground truth for clear and intelligible speech in this study was defined by listener derived ratings of naturalness and intelligibility. Traditionally speech is assessed by a clinician, who listens to a patient’ s speech and makes a subjective judgment about performance. Our study found that experts agreed on 61-70% of speech ratings. This considerable variability in subjective perception and evaluation of speech provides further motivation for pursuing objective and automated tools in its stead.

Speech is an expression of neurodegeneration and is therefore suitable for monitoring change, it is easy to elicit, it is repeatable, has multiple forms, there are elements that can be used across languages and its stability is known in some contexts. The benefits of being able to communicate effectively are also clear. There is consensus that maintaining clear and intelligible speech is vital for preserving quality of life. Further, its measurement is important, with patient advocacy groups consulting through the Food and Drug Administration (FDA) Externally-led Patient-Focused Drug Development Meeting on Friedreich Ataxia held in 2017 where more than two-thirds of patients noted that they are concerned about their communication ^25^. This provides strong rationale for trial outcome measures to include evaluation of speech. Moreover, we illustrated the longitudinal variation of expert raters’ assessments for the speech scoring assessments (naturalness and intelligibility), plotting those along aggregate acoustic scores in the form of the first three principal components. Aggregate acoustic scores appear sufficiently sensitive to capture longitudinal changes and may be useful to monitor speech assessment (naturalness, intelligibility) and FRDA symptoms assessment (FARS, FAIS) changes with respect to a specific participant’ s baseline scores.

The study is limited by several factors. Firstly, our sample was collated from a clinical cohort that was missing some data points across the 10-year period meaning not all participants provided all speech samples at each time point, nor did all participants provide samples each year. Secondly, we relied on subjective clinician or consumer rated measures of function and attempted to map them to objective markers of cognitive-linguistic function (acoustic measures). This method is acceptable when recognizing deficits, but not ideal for quantifying the size of change that can occur over time. Listener ratings are susceptible to poor inter/intra rater reliability and are not suitable for repeated application in the trial context ^3^. Despite these drawbacks, the human ear (especially when trained) can detect degrees of ‘abnormality’, and these were used to produce the multi-parameter acoustic features within this study.

## Data Availability

All data produced in the present manuscript are restricted for clinical investigation by the study team and are not publicly available at this time.

## Acknowledgements

Adam Vogel was supported by a National Health and Medical Research Council, Australia (Fellowship IDs 1082910 and 1135683) and the Australia Research Council (FT220100253). The study was supported at its early stages by the Wellcome Trust through Centre Grant No. 098461/Z/12/Z from the University of Oxford Sleep & Circadian Neuroscience Institute (SCNi). It was also supported by Health Data Research UK, which receives funding from HDR UK Ltd (HDR-5012) funded by the UK Medical Research Council, Engineering and Physical Sciences Research Council, Economic and Social Research Council, Department of Health and Social Care (England), Chief Scientist Office of the Scottish Government Health and Social Care Directorates, Health and Social Care Research and Development Division (Welsh Government), Public Health Agency (Northern Ireland), the British Heart Foundation, and the Wellcome Trust. The funders had no role in the study or the decision to submit this work to be considered for publication. Authors were not precluded from accessing data in the study, and they accept responsibility to submit for publication.

## FIGURES

**Fig 1**: Overview of the timing of successive participant visits in terms of the actual dates (a) and assessments relative to baseline visit (b).

**Fig. 2**: Out of sample performance for assessing a) intelligibility, b) naturalness, c) FAIS, d) FARS. Mean Absolute Error (MAE) was computed using 10-fold CV with 100 iterations for statistical confidence. Error bars represent the standard deviation around the quoted mean. FS = Feature Selection.

**Fig. 3**: Longitudinal variability of the four scales for participants who had at least four visits (although the scales were not necessarily available for all their visits). Indicative examples of longitudinal variation of intelligibility and naturalness and the aggregate acoustic information presented using the first three principal components. a) intelligibility, b) naturalness, c) PCA features, d) FARS, e) FAIS. Note: To facilitate visualization we have introduced slight jitter on the x-axis so that the markers do not completely overlap at the time of assessment. PCA feature subset based on voice measures only and did not include measures of timing due to inadequate tokens.

