## Supplementary material for "Clinically meaningful metrics of speech in neurodegenerative disease: Quantification of speech intelligibility and naturalness in ataxia": Supplementary Materials 29MARCH2023.docx

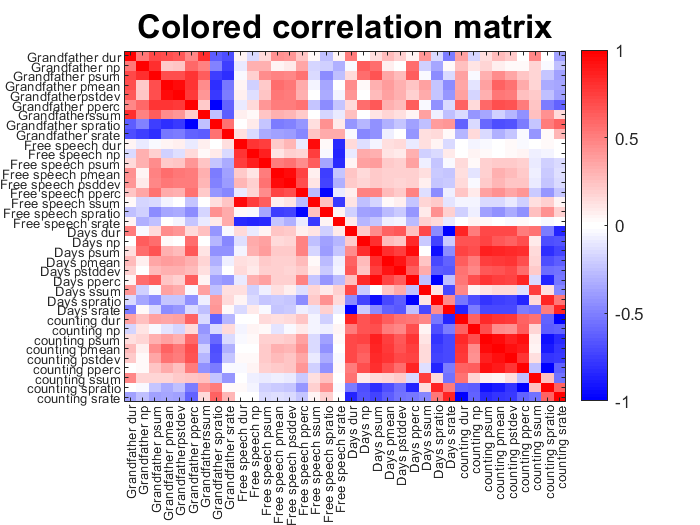

**Fig. S1**: Correlogram to visualize the internal correlations of the 36 timing measures.

Blue indicates that the two timing measures are strongly negatively correlated and red indicates strong positive correlation, whereas white suggests that the two timing measures are not correlated. See additional excel sheet for all correlations across measures.

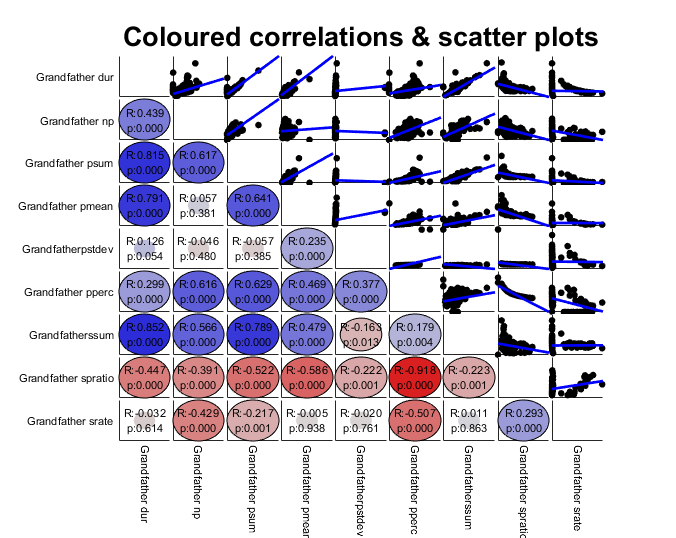

**Fig. S2**: Investigating pairwise statistical associations for the 9 acoustic measures extracted from the reading passage. The part above the long diagonal presents the scatter plots for the pairwise associations along with the best fit line (in blue color); below the long diagonal we illustrate the actual statistical correlations (R values) and statistical significance (p-values).

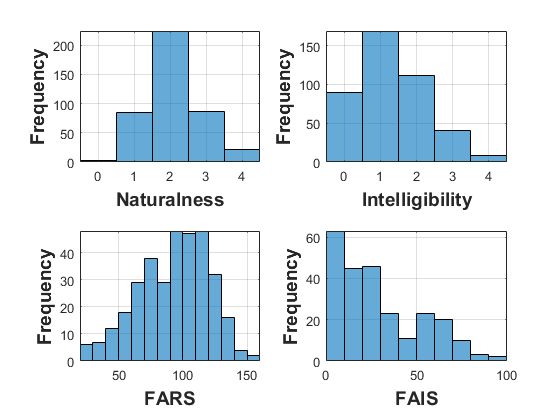

**Fig. S3**: Histogram with the distributions for the four clinical scales used in the study.

Table S1: Agreement matrix to indicate inter-rater variability for the intelligibility scores.

|  | **Rater 2** | | | | | |
| --- | --- | --- | --- | --- | --- | --- |
| **Rater 1** |  | ${\hat{\boldsymbol{y}}}_{\boldsymbol{0,R}\boldsymbol{2}}$ | ${\hat{\boldsymbol{y}}}_{\boldsymbol{1,R}\boldsymbol{2}}$ | ${\hat{\boldsymbol{y}}}_{\boldsymbol{2,R}\boldsymbol{2}}$ | ${\hat{\boldsymbol{y}}}_{\boldsymbol{3,R}\boldsymbol{2}}$ | ${\hat{\boldsymbol{y}}}_{\boldsymbol{4,R}\boldsymbol{2}}$ |
|  | ${\hat{\boldsymbol{y}}}_{\boldsymbol{0,R}\boldsymbol{1}}$ | **54** | 40 | 0 | 0 | 0 |
|  | ${\hat{\boldsymbol{y}}}_{\boldsymbol{1,R}\boldsymbol{1}}$ | 24 | **108** | 36 | 0 | 0 |
|  | ${\hat{\boldsymbol{y}}}_{\boldsymbol{2,R}\boldsymbol{1}}$ | 0 | 20 | **49** | 6 | 0 |
|  | ${\hat{\boldsymbol{y}}}_{\boldsymbol{3,R}\boldsymbol{1}}$ | 0 | 0 | 15 | **16** | 0 |
|  | ${\hat{\boldsymbol{y}}}_{\boldsymbol{4,R}\boldsymbol{1}}$ | 0 | 0 | 1 | 5 | **6** |

Intelligibility scores are scalar values in the range 0…4. Entries in the long diagonal suggest the two raters agreed in their assessment.

Table S2: Agreement matrix to indicate inter-rater variability for the naturalness scores.

|  | **Rater 2** | | | | | |
| --- | --- | --- | --- | --- | --- | --- |
| **Rater 1** |  | ${\hat{\boldsymbol{y}}}_{\boldsymbol{0,R}\boldsymbol{2}}$ | ${\hat{\boldsymbol{y}}}_{\boldsymbol{1,R}\boldsymbol{2}}$ | ${\hat{\boldsymbol{y}}}_{\boldsymbol{2,R}\boldsymbol{2}}$ | ${\hat{\boldsymbol{y}}}_{\boldsymbol{3,R}\boldsymbol{2}}$ | ${\hat{\boldsymbol{y}}}_{\boldsymbol{4,R}\boldsymbol{2}}$ |
|  | ${\hat{\boldsymbol{y}}}_{\boldsymbol{0,R}\boldsymbol{1}}$ | **2** | 4 | 0 | 0 | 0 |
|  | ${\hat{\boldsymbol{y}}}_{\boldsymbol{1,R}\boldsymbol{1}}$ | 4 | **48** | 19 | 0 | 0 |
|  | ${\hat{\boldsymbol{y}}}_{\boldsymbol{2,R}\boldsymbol{1}}$ | 0 | 29 | **139** | 20 | 0 |
|  | ${\hat{\boldsymbol{y}}}_{\boldsymbol{3,R}\boldsymbol{1}}$ | 0 | 1 | 24 | **60** | 5 |
|  | ${\hat{\boldsymbol{y}}}_{\boldsymbol{4,R}\boldsymbol{1}}$ | 0 | 0 | 0 | 8 | **17** |

Naturalness scores are scalar values in the range 0…4. Entries in the long diagonal suggest the two raters agreed in their assessment.

Additional context for speech as a marker of symptom change in neurodegenerative disease.

For many years it has been possible to record speech for offline analyses. However the expense and quality has limited the extent to which recorded speech could be used to improve clinical decision making or contribute to brain-behavior models of disease. Recent improvements in digital technology, including microphone design, computational power and signal processing, allow for more effective and usable tools for the collection of speech samples in both clinical and remote contexts with a fidelity sufficient to allow both subjective and objective analyses. This facility also allows rapid aggregation of the data from healthy individuals as well as individuals with brain disease or injury, thereby improving methods for the quantification of speech characteristics and classification of disordered speech. Such methods also allow sampling of speech to take place over shorter retest intervals, for example from annual to weekly clinical assessments, across different contexts (during work or at home), as well as providing the means to control the aspects of speech assessed in these different contexts. These advances have allowed assessments and analysis of speech to guide decisions about the presence ^1-3^ and progression ^4^ of brain diseases as well as the response of brain diseases to behavioral and pharmacotherapies ^5,6^.
